# MATHEMATICAL MODELING FOR TRANSMISSIBILITY OF COVID-19 VIA MOTORCYCLES

**DOI:** 10.1101/2020.04.18.20070797

**Authors:** Benard Okelo

## Abstract

In this paper, we present a mathematical model of trigonometric type for transmissibility and deaths as a result of COVID-19. In the model, we analyze the spread of COVID-19 by considering a new parameter, the motor-cycle as a means of public transport, which has not been considered in several other models for COVID-19. We use the mathematical model to predict the spread and deaths and we suggest strategies that can be put in place to prevent the spread caused by motorcycle as a means of public transport.

## 1. Introduction

The outbreak of COVID-19 was detected in December 2019 in mainland China with the city of Wuhan as the recognized epicenter(see [1]-[29] and the references therein). COVID-19 has since then been exported to other countries all over the world[7]. To describe and determine the dynamics of COVID-19, several mathematical models have been constructed by several researchers[2]-[6]. We realize that the insights that can be drawn from these models are discussed, especially as inputs for designing strategies to control the epidemics. Proposed model-based strategies on how to prevent the spread of the disease in local setting, such as during large social gatherings, are also discussed in details[8]-[13]. One of the parameters considered in most of the models is the exposure time which is an instrumental factor in spreading the disease[14]. With a basic reproduction number equal to 2, and 14-day infectious period, an infected person staying more than 9 hours in the event could infect other people [15]. As at now, there is an urgent need to develop a mathematical model to estimate the transmissibility and dynamic of the transmission of the virus[16]-[19]. Several researches [17], [18], [21], [22], [22] and [25] have focused on calculating the basic reproduction number *R*_0_ by using the serial intervals and intrinsic growth rate or using ordinary differential equations and Markov Chain Monte Carlo methods[23] and [24]. A lot of these researches have certain similar characteristics: They show that social distancing can significantly slow down the spread of the COVID-19; They stipulated that the effectiveness depends very much on the initial and boundary conditions of social distancing and the results strongly vary by orders of magnitude if the process is initiated a few days earlier or later or the effectiveness of the measure is only slightly higher or lower; The details of the studies are difficult to understand for many readers and therefore a widespread trust on the efficiency on the actual measures regarding the COVID-19 pandemic can not easily be derived from scientific literature. Results from [26] show that the effectiveness of social distancing is particularly great for pandemic with a basic reproductive number of *R*_0_ = 1.5 - 2.5. For the COVID-19 pandemic, a basic reproduction number *R*_0_ = 2.6 is estimated, with an uncertainty range from 1.5 to 3.5 as seen in [27]. *R*_0_ specifies how many new cases are caused by an existing case on average. With *R*_0_¿ 1, the virus spreads exponentially at times, but for *R*_0_ ¡1 the spread stops on its own [30]. A large number of studies have now calculated the number of basic reproductions, the incubation period and the duration of the infectiousness based on the number of cases from China [30]. However, the transmission through use of motorcycles as a means of public transport particularly in developing economies have not considered in the published models. The COVID-19 therefore is still a world pandemic and is currently a challenge for several countries all over the the world but worse for the developing countries. The goal of this paper is to present a mathematical model of trigonometric form for the spread of the COVID-19, where we concentrate on a new parameter called the use of motorcycles as a means of public transport to predict spread and number of deaths due to the COVID-19.

## 2. Materials and methods

The reported cases of COVID-19, were collected for the modelling study from the World Health Organization official website. The epidemic curve from a sample of 30 days is presented. Simulation methods and statistical analysis was done using MATLAB version 9.7. The fourth-order RungeKutta method, with tolerance set at 0.001, was used to perform curve fitting. Selected preliminary models of COVID-19, published in official webpages of academic/research institutions, as preprints, or as journal articles since the outbreak are reviewed. Important insights from the results of these models are discussed. Moreover, a model using a Susceptible-Exposed-Infected (SEI) framework is has been carefully reviewed to propose measures to prevent epidemics during large events, e.g., during parties or concerts with huge crowds. The parameter values used in this model are based on the known information about COVID-19. The model is represented by a system of instantaneous and noninstantaneous differential equations, and simulated using MATLAB version 9.7. The Differential equations are then reduced into a trigonometric form and later analysed. Motorcycles are used in most developing countries for public transport. In Kenya, they are called “Boda Boda”, while in Nigeria they are commonly known as “Okada.” Most people in the cities, towns and even in rural areas prefer motorcycle because they are faster, relatively cheap and can be used to avoid traffic jam in roads. Despite their usefulness, motorcycles contributes immensely to the spread of COVID-19 because the operators do not follow the traffic rules. For instance, they don’t put masks and helmets, they carry excess passengers, they don’t wash their hands as required. Hence, social distancing is not taken care of. Due to these factors, we study the transmissibility of COVID-19 as a result of the risk factors from the motorcycle operators.

## 3. Derivation of the mathematical model

We consider the use of motorcycles as a parameter presented in form of differential equations with instantaneous impulses and we compare it with the case of noninstantaneous impulses.

### Case I. Instantaneous impulses

Let an increasing sequence of points 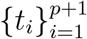 be given such that 0 ≤ *t*_*i*_ *< t*_*i*+1_, *i* = 1, 2, …, *p*, lim_*k*→∞_ *t*_*k*_ = ∞ Let *t*_0_ ∈ *R*^*n*^ be a given arbitrary point. Without loss of generality we assume that *t*_0_ ∈ [0, *t*_1_). Consider the Initial Value Problem (IVP) for the nonlinear Instantaneous Impulsive Differential equation (IDE)

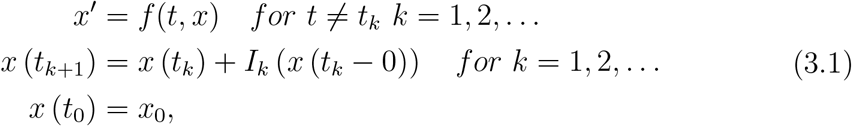

where *x, x*_0_ ∈ ℝ^*n*^, *f* : [*t*_0_, ∞) *×*ℝ^*n*^ → ℝ^*n*^, *I*_*k*_ : ℝ^*n*^ → ℝ^*n*^, (*k* = 1, 2, 3, …). The points *t*_*k*_, *k* = 1, 2, … are called points of instantaneous impulses and the functions *I*_*k*_(*x*), *k* = 1, 2, … are called instantaneous impulsive functions. The solution *x* (*t*; *t*_0_, *x*_0_) in the general case is a piecewise continuous function which is satisfying the integral equation

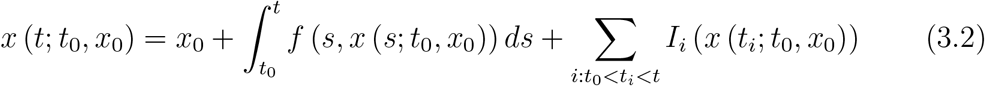

### Case II. Noninstantaneous impulses

Let two increasing finite sequences of points 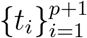 and 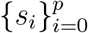 be given such that *s*_0_ = 0 *< t*_*i*_ ≤ *s*_*i*_ *< t*_*i*+1_, *i* = 1, 2, 3, …, *p* and points *t*_0_, *T* ∈ ℝ_+_ are given such that *s*_0_ = 0 *< t*_0_ *< t*_1_, *t*_*p*_ *< T ≤ t*_*p*+1_, *p* is a natural number. Consider the Initial Value Problem(IVP) for the nonlinear Noninstantaneous Impulsive Differential Equation(NIDE)

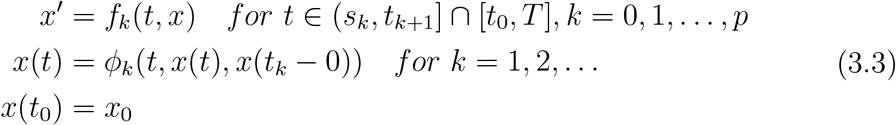

where *x, x*_0_ ∈ ℝ, *f*_*k*_ : [*s*_*k*_, *t*_*k*+1_] ∩ [*t*_0_, *T*] *×*ℝ → ℝ, *k* = 0, 1, 2, …, *p, ϕ*_*k*_ : [*t*_*k*_, *s*_*k*_] *×*ℝ *×* ℝ → ℝ, *k* = 1, 2, 3, … The intervals (*s*_*k*_, *t*_*k*+1_], *k* = 1, 2, … are called intervals of noninstantaneous impulse and the function *ϕ*_*k*_(*t, x, y*), *k* = 1, 2, …, are called noninstantaneous impulsive function. In the special case *s*_*k*_ = *t*_*k*+1_, *k* = 0, 1, 2, … each interval of noninstantaneous impulses is reduced to a point, and the problem in Equation 3.3 is reduced to an IVP for an IDE in Equation 3.1 with points of impulses *t*_*k*_ and impulsive functions *x* (*t*_*k*_ + 0) = *I*_*k*_ (*x* (*t*_*k*_ − 0)) ≡ *ϕ*_*k*_ (*t*_*k*_, *x* (*t*_*k*_ − 0), *x* (*t*_*k*_ − 0)) − *x* (*t*_*k*_ − 0) Let *k* ≥ 0 be a given integer, *τ* ∈ [*t*_*k*_, *s*_*k*_) be a given point and consider the corresponding IVP for ODE

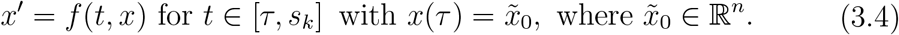

We give a brief description of the solution of IVP for NIDE in Equation 3.4 The solution *x* (*t*; *t*_0_, *x*_0_) of Equation 3.3 is given by

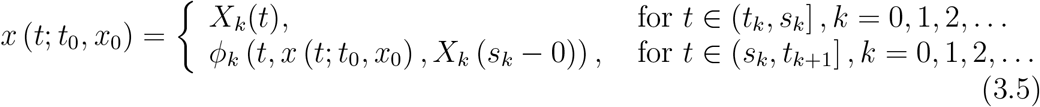

where; on the interval [*t*_0_, *s*_0_] the solution coincides with *X*_0_(*t*) which is the solution of IVP for ODE in Equation 3.4 for *τ* = *t*_0_, *k* = 0 and 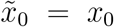 on the interval (*s*_0_, *t*_1_] the solution *x* (*t*; *t*_0_, *x*_0_) satisfies the equation *x* (*t*; *t*_0_, *x*_0_) = *ϕ*_0_ (*t, x* (*t*; *t*_0_, *x*_0_), *X*_0_ (*s*_0_ − 0)); on the interval (*t*_1_, *s*_1_] the solution coincides with *X*_1_(*t*) which is the solution of IVP for ODE in Equation 3.4 for *τ* = *t*_1_, *k* = 1 and 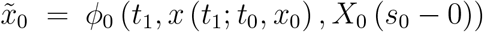; on the interval (*s*_1_, *t*_2_] the solution *x* (*t*; *t*_0_, *x*_0_) satisfies the equation *x* (*t*; *t*_0_, *x*_0_) = *ϕ*_1_ (*t, x* (*t*; *t*_0_, *x*_0_), *X*_1_ (*s*_1_ − 0)); on the interval (*t*_2_, *s*_2_] the solution coincides with *X*_2_(*t*) which is the solution of IVP for ODE in Equation 3.4 for *τ* = *t*_2_, *k* = 2 and 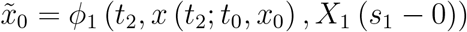; and so on. Also, the solution *x* (*t*; *t*_0_, *x*_0_), *t ≥ t*_0_ of Equation 3.3 satisfies the following system of integral and algebraic equations

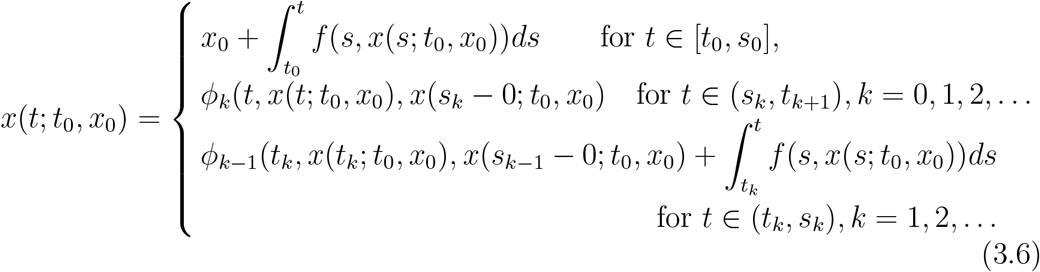

**Example 3.1**. Consider the IVP for the scalar NIDE

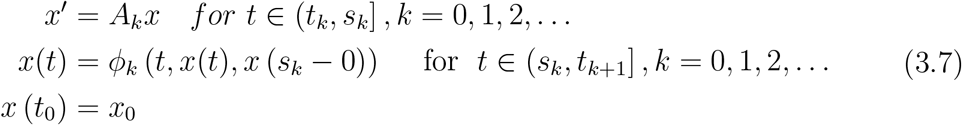

where *x, x*_0_ ∈ ℝ, *A*_*k*_, *k* = 0, 1, 2, …, are constants. The solution of Equation 3.7 is given by

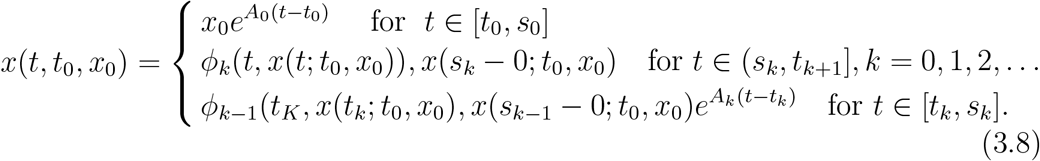

Now, we consider a number of cases:

**Case 1**. Let *A*_*k*_ = *A, ϕ*_*k*_(*t, x, y*) = *a*_*k*_(*t*)*x*^2^*y, a*_*k*_ : [*s*_*k*_, *t*_*k*+1_] → ℝ*/{*0*}, k* = 0, 1, 2, … and *x*_0_ ≠ 0. since the nontrivial solution of *x* = *a*_*k*_(*t*)*x*^2^*y* for any *y*∈ ℝ, *y* ≠ 0 is 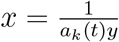, then the solution of NIDE in Equation 3.7 is given by

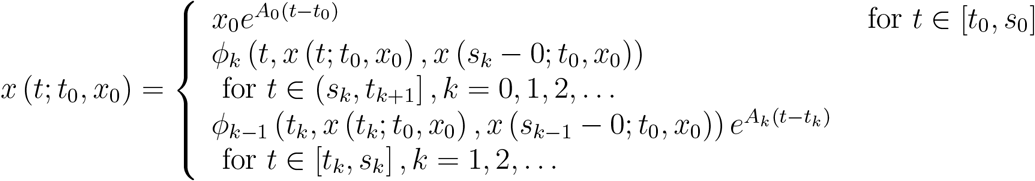

A particular case where *A* = 1, *s*_*k*_ = 2*k*− 1, *t*_*k*_ = 2*k, a*_*k*_(*t*) = *t, k* = 0, 1, 2 …, 5 has solutions for two different initial values *x*_0_ = 0.5 and *x*_0_ = 0.3.

**Case2**. Let *A*_*k*_ = *A, ϕ*_*k*_(*t, x, y*) = *a*_*k*_(*t*)*y, a*_*k*_ : [*s*_*k*_, *t*_*k*+1_] → ℝ, *k* = 0, 1, 2, 3, … The solution NIDE in Equation 3.7 is given by

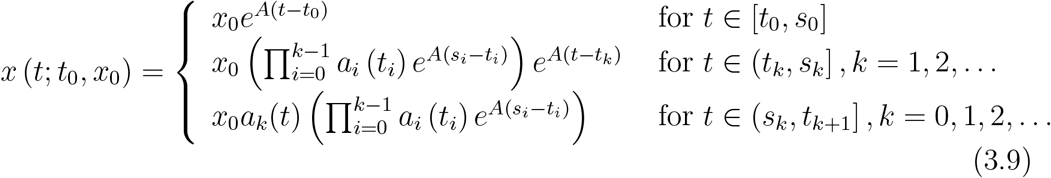

In the particular case *A* = −1, and *t*_*k*_ = 2*k, s*_*k*_ = 2*k*+1, *a*_*k*_(*t*) = *t* for *k* = 0, 1, 2, … we have the solutions for the different initial values *x*_0_ = 1, 0.5 and 0.2.

**Case 3**. Let *A* = 0 and *ϕ*_*k*_(*t, x, y*) = *a*_*k*_(*t*)*y, a*_*k*_ : [*t*_*k*_, *s*_*k*_] → ℝ, *k* = 1, 2, …, *p* The solution of NIDE in Equation 3.7 is given by

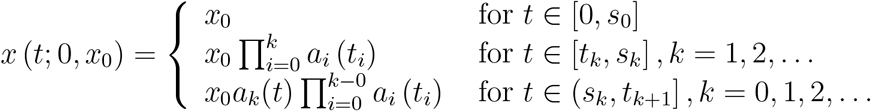

**Example 3.2**. Comparisons between the behavior motorcycle operators that leads to the spread of COVID-19 with the corresponding impulsive differential equation and the equation with non-instantaneous impulses are given in the following cases.

**I**. Ordinary Differential Equations. Consider the IVP for the scalar ODE

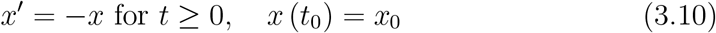

where *x*_0_ ∈ ℝ. The solution of Equation 3.10 is 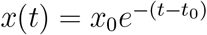 and it approaches 0 as *t* →∞.

**II**. Impulsive differential equations. Consider the IVP for the scalar Impulsive Differential Equation (IDE)

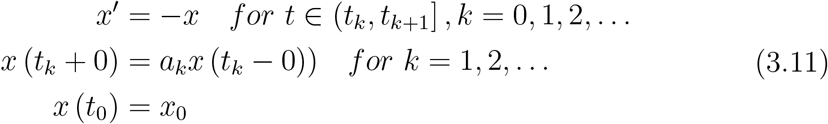

where *x*_0_ ∈ ℝ, *a*_*k*_, *k* = 1, 2,… *· k* are constants. The solution of Equation 3.11 is given by 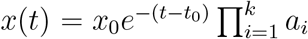 for *t* ∈ (*t*_*k*_, *t*_*k*+1_]. The behavior of the solution depends significantly on the amount of the impulse, i.e., the value of the constants *a*_*k*_.

**Case II.1**. If *a*_*k*_ = 1, *k* = 1, 2, …, then the problem in Equation 3.11 coincides with the IVP for ODE in Equation 3.10 and the solution approaches 0 as *t* → ∞.

**Case II.2**. If there exists a natural number *k* : *a*_*k*_ = 0, then the solution of Equation 3.11 is zero for *t > t*_*k*_, i.e., all solutions in spite of the initial value *x*_0_ coincide for *t > t*_*k*_.

**Case II.3**. If *a*_*k*_ = *e*^−*k*^, *t*_*k*_ = *k, k* = 1, 2, …, then the solution of the problem in Equation 3.11 is *x*(*t*) = *x*_0_*e*^*t*−*k*^ for *t* ∈ (*k, k* + 1] and the solution is a periodic function.

**Case II.4**. If *a*_*k*_ = *e*^−2*k*^, *t*_*k*_ = *k, k* = 1, 2, …, then the solution of the problem in Equation 3.11 is *x*(*t*) = *x*_0_*e*^*t*−2*k*^ for *t* ∈ (*k, k* + 1] and the solution being a piecewise continuous function approaches 0.

**Case II.5**. If *a*_*k*_ = *e*^−2*k*^, *t*_*k*_ = *k, k* = 1, 2, …, then the solution of the problem in Equation 3.11 is *x*(*t*) = *x*_0_*e*^*t*+0.5*k*^ for *t* ∈ (*k, k* + 1] and the solution is an unbounded function.

**III**. Differential equations with non-instantaneous impulses. Consider the IVP for the scalar NIDE

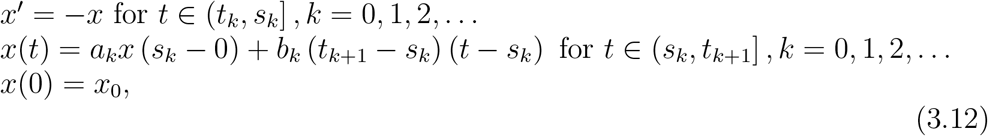

where *x*_0_ ∈ ℝ, *a*_*k*_, *k* = 0, 1, 2, … *k* are constants, *t*_*k*_ = *k, s*_*k*_ : *k < s*_*k*_ ≤ *k* + 1 for *k* = 0, 1, … Note in the case *t*_*k*+1_ = *s*_*k*_ the second equation of 3.12 is reduced to *x* (*t*_*k*_ + 0) = *a*_*k*_*x* (*t*_*k*_ − 0). We consider the case *t*_*k*+1_ ≠ *s*_*k*_. For the spontaneous transmission of COVID-19 by the motorcycles, the solution is given by

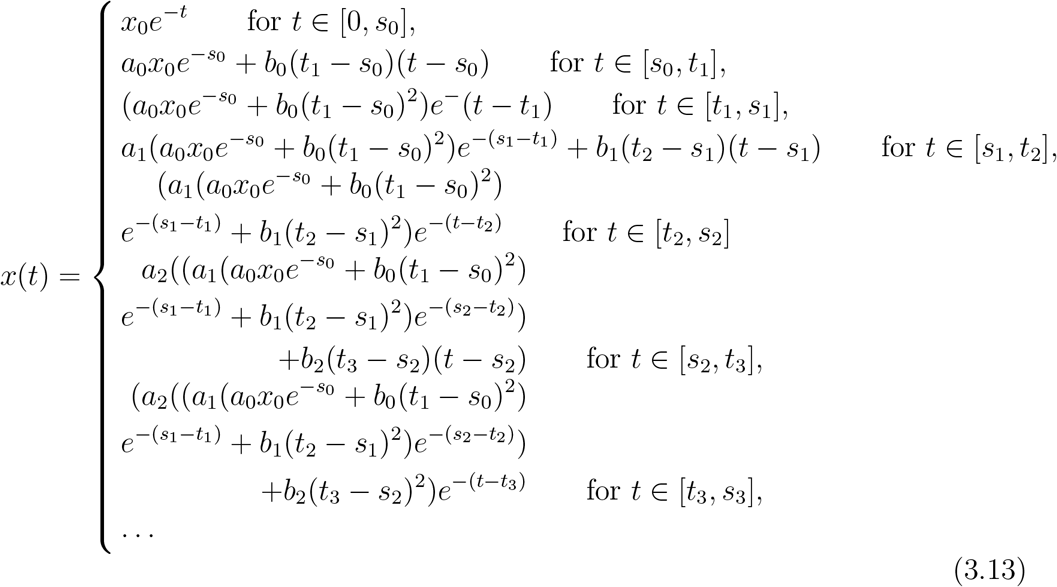

If *a*_*k*_ = 0, *k* = 1, 2, …, then the solution of (3.1.12) is given by

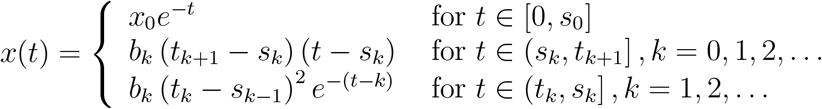

The solutions do not depend on the initial value *x*_0_ for *t > t*_1_

At this point, we consider the mathematical models with motorcycles as a parameter trigonometrically.

Consider Equation 3.13. With some complex and tedious computation, we can trigonometrically represent it as

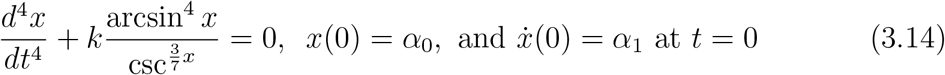

where *k* is a positive real number.

We note that the days in the period at which 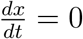 and 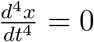 are points when COVID-19 is neither being transmitted nor spread and no deaths are realized. Now taking the following in account: At 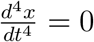 and 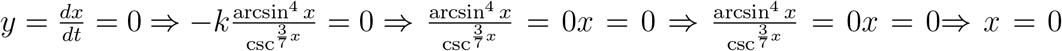 (as the only solution). The day considered to be the point (0, 0) is the maximal day point which is stable as per the fourth-order RungeKutta method, with tolerance set at 0.001, was used to perform curve fitting. Since Equation 3.14 is positive definite while 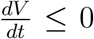, the origin is Lyapunov stable with a trajectory curve 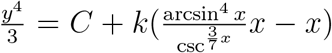. Considering the maximal point and the initial conditions of Equation 3.14 we get

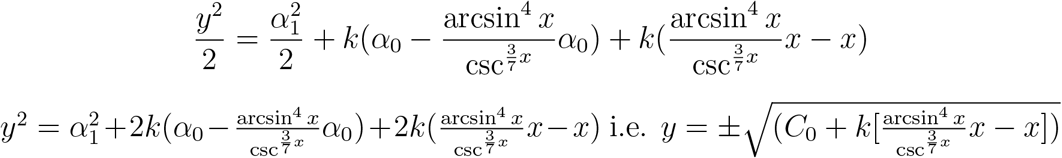

From Figure 1 below, considering different transmission levels i.e. *C*_0_ = 0, *C*_0_ = 5 and *C*_0_ = 10 and putting *k* = 1, the curve in the phase plane has symmetry in the *x* axis only.

**Figure 1.**
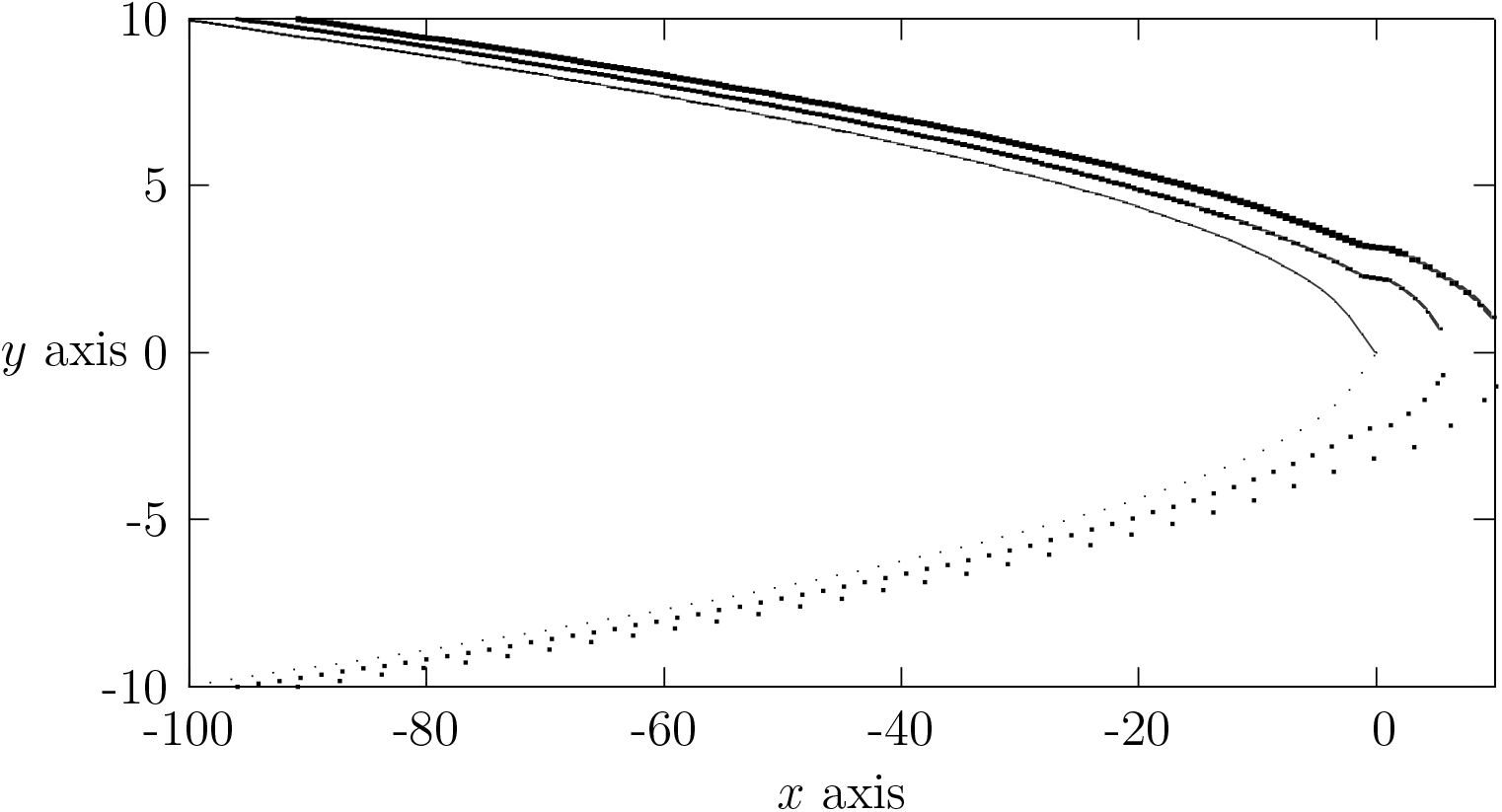
COVID-19 transmissibility at the maximal points

From

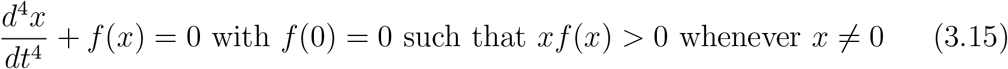

It can be shown that a Lyapunov function *V* (*x, y*) for the Equation 3.15 is given by

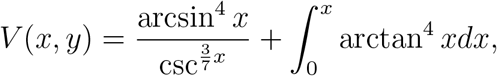

and the origin (0, 0) is a stable equilibrium point of the system of differential Equation 3.15. In fact Equation 3.14 is but a particular case of Equation 3.15. One easily deduces that though the origin (0, 0) is stable, it is not asymptotically stable.

Data was collected over 30 days with regard to positivity indices from COVID-19 cases and questions were asked from the respondents with regard to use of motorcycles a public means of transport in different countries all over the world.

This leads to the following results:

| t | x when $\Phi = 1.7$<br>and $\Psi = 0.001$ | x when $\Phi = \frac{3}{7}$<br>and $\Psi = \frac{3}{7}$ | x when $\Phi = 1.5$<br>and $\Psi = 1.5$ |
| --- | --- | --- | --- |
| Day | 1.7 | 1.7 | 1.7 |
| 1 | 0.35 | 0.35 | 0.35 |
| 2 | 0.432593 | 0.471592 | 0.473409 |
| 3 | 0.521135 | 0.54544 | 0.557654 |
| 4 | 0.545426 | 0.558432 | 0.565432 |
| 5 | 0.567833 | 0.508191 | 0.509875 |
| 6 | 0.445835 | 0.403806 | 0.418790 |
| 7 | 0.3789038 | 0.262878 | 0.560165 |
| 8 | 0.128916 | 0.104905 | 0.1020876 |
| 9 | -0.0800323 | -0.0567865 | -0.098701 |
| 10 | -0.288008 | -0.220356 | -0.2398765 |
| 11 | -0.511782 | -0.396754 | -0.4240985 |
| 12 | -0.779739 | -0.609934 | -0.648768 |
| 13 | -1.13535 | -0.898417 | -0.949256 |
| 14 | -1.63076 | -1.31534 | -1.98100 |
| 15 | -2.31625 | -1.91814 | -2.06576 |
| 16 | -3.22819 | -2.74816 | -2.87880 |
| 17 | -4.38406 | -3.82302 | -3.94118 |
| 18 | -5.78896 | -5.14714 | -5.28242 |
| 19 | -7.44377 | -6.72119 | -6.87359 |
| 20 | -9.34856 | -8.54524 | -8.71475 |
| 21 | -11.5034 | -10.6193 | -10.8059 |
| 22 | -13.9082 | -12.9433 | -13.1471 |
| 23 | -16.563 | -15.9074 | -15.7382 |
| 24 | -19.40098 | -28.3414 | -18.5794 |
| 25 | -22.60943 | -21.4155 | -21.6706 |
| 26 | -26.0273 | -27.7395 | -25.0117 |
| 27 | -41.6821 | -29.3136 | -28.6029 |
| 28 | -33.5869 | -33.1399 | -32.4441 |
| 29 | -37.7417 | -36.2887 | -26.5352 |
| 30 | -32.1333 | -30.5357 | -30.8887 |

The curve in Figure 2 shows that the COVID-19 is spread through the use of motorcycle and the rate of infection is continuous. The curve reaches the peak but the rate of reduction in terms of death and infections is not rapid. Therefore, the pandemic might take longer than expected for it to be contained and to be declared not to be a pandemic anymore.

**Figure 2.**
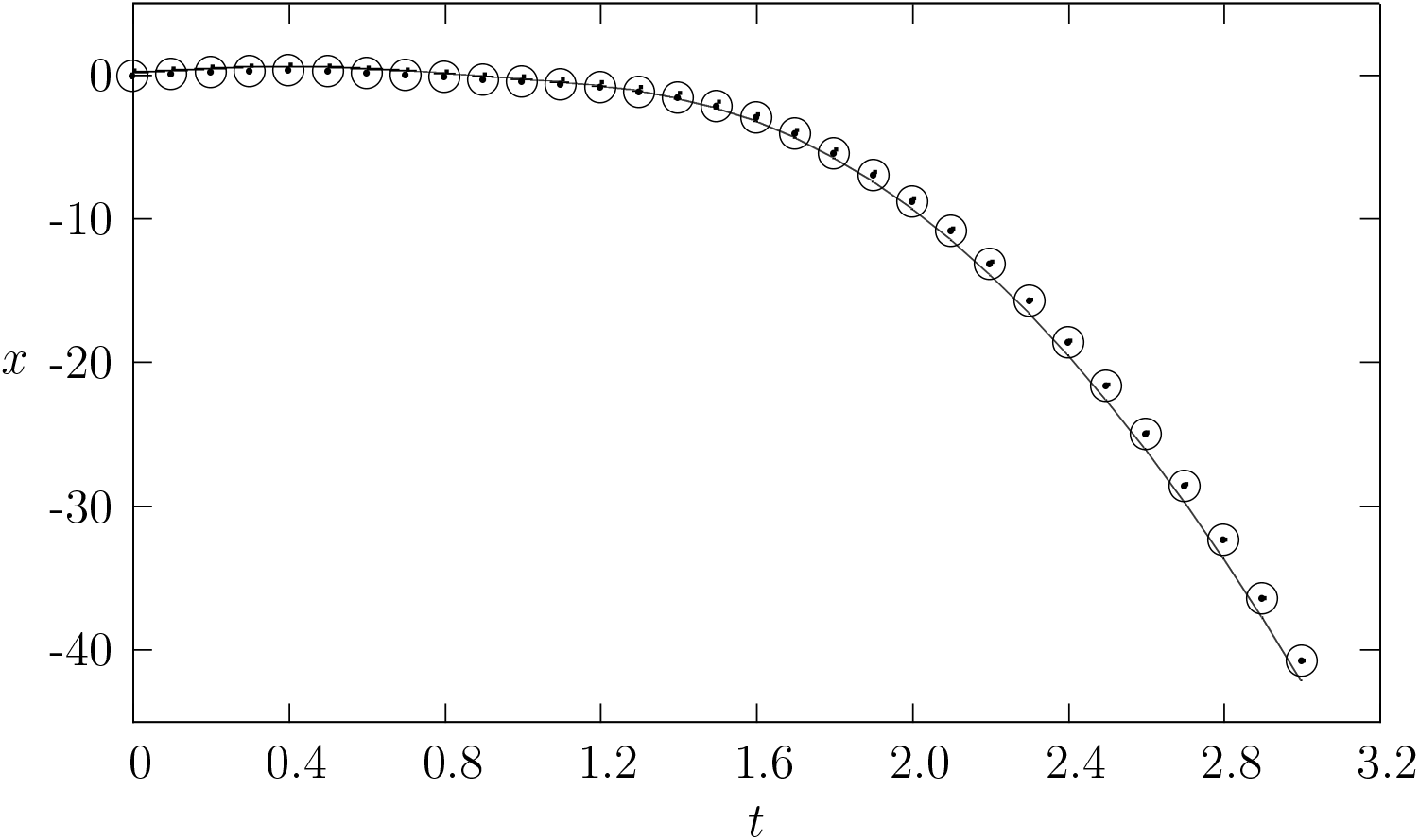
The curve of COVID-19 transmissibility over thirty day period through motorcycle operations.

## 4. Conclusion

The results from this study is in agreement with the recommendation of World Health Organization [22] that it is still possible to interrupt virus spread, provided that countries put in place strong measures to detect disease early, for example, development of rapid diagnostic tests, and increasing effectiveness of passenger screening in airports in which thermal screening for COVID-19 infection is estimated using simulation to be 46 percent. Moreover, motorcycle operators are not indispensable when it comes to observing recommendation of World Health Organization and therefore if they adhere strictly to these recommendations, there will be a tremendous decrease in new infections and consequently reduces deaths due to COVID-19.

## Data Availability

Data was simulated from sotwares

